# Polygenic risk affects the penetrance of monogenic kidney disease

**DOI:** 10.1101/2023.05.07.23289614

**Authors:** Atlas Khan, Ning Shang, Jordan G. Nestor, Chunhua Weng, George Hripcsak, Peter C. Harris, Ali G. Gharavi, Krzysztof Kiryluk

## Abstract

**Background:** Chronic kidney disease (CKD) is a genetically complex disease determined by an interplay of monogenic, polygenic, and environmental risks. Most forms of monogenic kidney diseases have incomplete penetrance and variable expressivity. It is presently unknown if some of the variability in penetrance can be attributed to polygenic factors.

**Methods:** Using the UK Biobank (N=469,835 participants) and the All of Us (N=98,622 participants) datasets, we examined two most common forms of monogenic kidney disorders, autosomal dominant polycystic kidney disease (ADPKD) caused by deleterious variants in the *PKD1* or *PKD2* genes, and COL4A-associated nephropathy (COL4A-AN caused by deleterious variants in *COL4A3*, *COL4A4*, or *COL4A5* genes). We used the eMERGE-III electronic CKD phenotype to define cases (estimated glomerular filtration rate (eGFR) <60 mL/min/1.73m2 or kidney failure) and controls (eGFR >90 mL/min/1.73m2 in the absence of kidney disease diagnoses). The effects of the genome-wide polygenic score (GPS) for CKD were tested in monogenic variant carriers and non-carriers using logistic regression controlling for age, sex, diabetes, and genetic ancestry.

**Results:** As expected, the carriers of known pathogenic and rare predicted loss-of-function variants in *PKD1* or *PKD2* had a high risk of CKD (OR_meta=_17.1, 95% CI: 11.1-26.4, P=1.8E-37). The GPS was comparably predictive of CKD in both ADPKD variant carriers (OR_meta=_2.28 per SD, 95%CI: 1.55-3.37, P=2.6E-05) and non-carriers (OR_meta=_1.72 per SD, 95% CI=1.69-1.76, P< E-300) independent of age, sex, diabetes, and genetic ancestry. Compared to the middle tertile of the GPS distribution for non-carriers, ADPKD variant carriers in the top tertile had a 54-fold increased risk of CKD, while ADPKD variant carriers in the bottom tertile had only a 3-fold increased risk of CKD. Similarly, the GPS was predictive of CKD in both COL4-AN variant carriers (OR_meta=_1.78, 95% CI=1.22-2.58, P=2.38E-03) and non-carriers (OR =1.70, 95%CI: 1.68-1.73 P<E-300). The carriers in the top tertile of the GPS had a 2.5-fold higher risk of CKD while the risk for carriers in the bottom tertile was similar to the middle tertile of non-carriers.

**Conclusions:** Variable penetrance of kidney disease in ADPKD and COL4-AN is partially explained by differences in polygenic risk profiles. Accounting for polygenic factors has the potential to improve risk stratification in monogenic kidney disease and may have implications for genetic counseling.

## INTRODUCTION

Common complex traits are determined by a combination of genetic and environmental risk factors. A small subset of common human diseases is caused by rare monogenic variants with relatively large effects that cause disease by disrupting a specific disease-related pathway^1, 2^. However, monogenic disease variants typically have incomplete penetrance that is often attributable to environmental and other inherited factors. Genome-wide polygenic scores (GPS) have emerged as a powerful approach to quantifying the contribution of polygenic effects^3–24^. Recent studies suggested that such scores could, to some degree, explain variable penetrance of several monogenic disorders, including familial hypercholesterolemia, hereditary breast, and ovarian cancer, and Lynch syndrome^25^. However, the interplay of monogenic and polygenic risk has not been previously studied in the context of kidney disease.

Chronic kidney disease (CKD) is a common condition that affects more than 10% of the population worldwide ^26^. CKD represents a genetically complex and highly heterogeneous phenotype. Monogenic disorders account for up to 9.3% of all-cause CKD^27^ with autosomal dominant polycystic kidney disease (ADPKD) and Alport syndrome, Thin Basement Membrane Disease, and Hereditary Nephritis, collectively known as collagen type IV-alpha-associated nephropathies (*COL4A*-AN) representing the most common forms of monogenic kidney diseases. ADPKD is caused by dominant mutations in the *PKD1* gene on chromosome 16 or the *PKD2* gene on chromosome 4. The disease affects all ancestral groups with an overall prevalence of approximately 1 in 1,000^28^. The second most common group of inherited nephropathies, COL4A-AN, are caused by mutations in *COL4A3*, *COL4A4*, or *COL4A5* genes. COL4A-AN is characterized by glomerular basement defects manifesting with hematuria and renal dysfunction. Biallelic inheritance causes Alport syndrome, a rare and more severe disease characterized by hematuria, early-onset kidney failure, and deafness. However, monoallelic carriers of pathogenic variants also have a higher risk of hematuria and CKD. The penetrance of both ADPKD and COL4A-AN is highly variable, even within the same pedigrees. In this study, we hypothesize that polygenic background may partially explain the observed variability in the penetrance of these disorders.

We hypothesize that while monogenic variants exert large effects by perturbing an essential disease pathway, polygenic risk factors can either ameliorate or exacerbate this effect by altering a broader array of mechanisms related to CKD. We have previously developed a GPS for CKD with validated performance across diverse ancestries^29^. Here, we test if the GPS modifies the risk of CKD among carriers of pathogenic ADPKD and COL4A-AN variants through combined analysis of exome/genome sequencing, SNP microarray, and electronic health record (EHR) data for a total of 568,457 participants from the UK Biobank and the All of Us study.

## METHODS

### Study design

This cross-sectional study involves a combined analysis of the UK Biobank (UKBB) and All of Us cohorts (AoU) cohorts. All participants provided informed consent to participate in genetic studies. Each cohort was first analyzed separately, and cohort-specific results were combined using fixed effects meta-analysis.

### UK Biobank (UKBB)

The UK Biobank is a longitudinal cohort of individuals ages 40-69 years at enrollment, recruited between 2006 and 2010 across the United Kingdom^30^. The individuals recruited to UKBB signed an electronic consent to allow a broad sharing of their anonymized data for health-related research. UKBB generated and released SNP microarray, exome sequence, and structured EHR data for 469,835 participants. The cohort is 54% female, with a mean age of 57 years, and the composition is 94% Europeans, 2% West or Southeast Asians, and 2% African ancestry by self-report^30^ (**Table 1**).

**Table 1:**
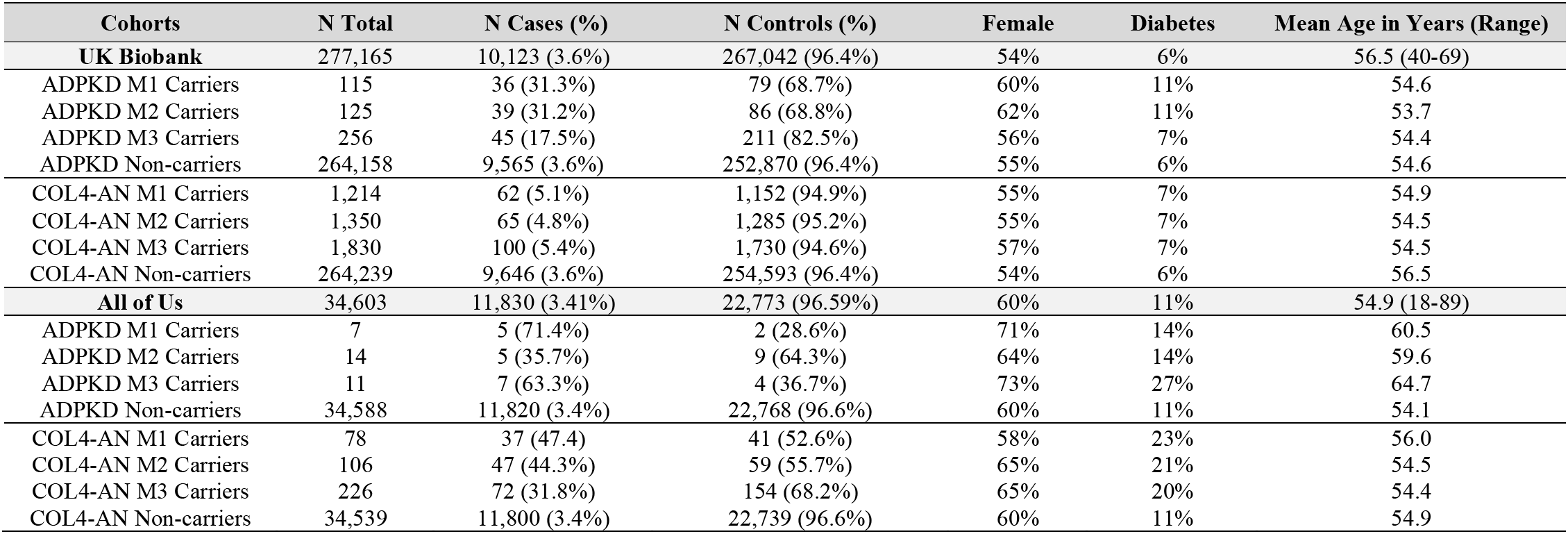
M1, M2, and M3 variant carriers and their characteristics in the UKBB and AoU datasets. The counts include only individuals with a valid phenotype (case/control) label that was included in the analyses. Note that the non-carrier group is common to all three variant models and excludes any M1, M2, and M3 variant carriers.

| Cohorts | N Total | N Cases (%) | N Controls (%) | Female | Diabetes | Mean Age in Years (Range) |
| --- | --- | --- | --- | --- | --- | --- |
| **UK Biobank** | 277,165 | 10,123 (3.6%) | 267,042 (96.4%) | 54% | 6% | 56.5 (40-69) |
| ADPKD M1 Carriers | 115 | 36 (31.3%) | 79 (68.7%) | 60% | 11% | 54.6 |
| ADPKD M2 Carriers | 125 | 39 (31.2%) | 86 (68.8%) | 62% | 11% | 53.7 |
| ADPKD M3 Carriers | 256 | 45 (17.5%) | 211 (82.5%) | 56% | 7% | 54.4 |
| ADPKD Non-carriers | 264,158 | 9,565 (3.6%) | 252,870 (96.4%) | 55% | 6% | 54.6 |
| COL4-AN M1 Carriers | 1,214 | 62 (5.1%) | 1,152 (94.9%) | 55% | 7% | 54.9 |
| COL4-AN M2 Carriers | 1,350 | 65 (4.8%) | 1,285 (95.2%) | 55% | 7% | 54.5 |
| COL4-AN M3 Carriers | 1,830 | 100 (5.4%) | 1,730 (94.6%) | 57% | 7% | 54.5 |
| COL4-AN Non-carriers | 264,239 | 9,646 (3.6%) | 254,593 (96.4%) | 54% | 6% | 56.5 |
| **All of Us** | 34,603 | 11,830 (3.41%) | 22,773 (96.59%) | 60% | 11% | 54.9 (18-89) |
| ADPKD M1 Carriers | 7 | 5 (71.4%) | 2 (28.6%) | 71% | 14% | 60.5 |
| ADPKD M2 Carriers | 14 | 5 (35.7%) | 9 (64.3%) | 64% | 14% | 59.6 |
| ADPKD M3 Carriers | 11 | 7 (63.3%) | 4 (36.7%) | 73% | 27% | 64.7 |
| ADPKD Non-carriers | 34,588 | 11,820 (3.4%) | 22,768 (96.6%) | 60% | 11% | 54.1 |
| COL4-AN M1 Carriers | 78 | 37 (47.4) | 41 (52.6%) | 58% | 23% | 56.0 |
| COL4-AN M2 Carriers | 106 | 47 (44.3%) | 59 (55.7%) | 65% | 21% | 54.5 |
| COL4-AN M3 Carriers | 226 | 72 (31.8%) | 154 (68.2%) | 65% | 20% | 54.4 |
| COL4-AN Non-carriers | 34,539 | 11,800 (3.4%) | 22,739 (96.6%) | 60% | 11% | 54.9 |

#### SNP Microarray data

The details of the UKBB microarray genotyping, imputation, and quality control are available elsewhere^30^. Briefly, using the UKBB Axiom Array (N=438,427) and UK BiLEVE Axiom Array (N=49,950), a total of 488,377 participants have been genotyped for 805,426 overlapping markers. The 1000 Genomes, UK10K, and Haplotype Reference Consortium (HRC) reference panels were all used in conjunction to perform genome-wide imputation using IMPUTE2 software^31, 32^. We performed post-imputation quality control analyses as described in our previous work based on this dataset,^29^ retaining 9,233,643 common (i.e., Minor Allele Frequency (MAF) > 0.01), high-quality (imputation R^2^ > 0.80) variants for the purpose of GPS calculation.

#### Exome sequencing

The exome sequencing (ES) dataset was generated for N=469,835 UKBB participants as previously described^33, 34^. Briefly, ES was performed at the Regeneron Genetics Center using 75 base pair pared-end reads with 10 base pair index reads on the Illumina NovaSeq 6000; the reads were mapped to the Genome Reference Consortium Human Reference 38 (GRCh38) using the BWA-MEM command for each sample. WeCall was used to identify variants in gVCFs, which were then aggregated with GLnexus into a joint-genotyped and multi-sample project-level VCF (pVCF). SNV and indel genotypes called threshold read depth (DP) were less than 7 and 10, respectively. Subsequent variant-level filters include at least one homozygous variant carrier; or at least one heterozygous variant carrier with an allele balance greater than 0.15 for SNVs and 0.20 for indels^33, 34^. We accessed and analyzed the latest data through the UKBB Research Analysis Platform (RAP) on DNAnexus. For the purpose of this study, we applied additional variant-level filters that included genotype quality (GQ) >90, depth of coverage (DP) >10, and MAF less than or equal to 0.00001 for ADPKD and 0.001 for COL4A-AN variants in the UKBB and GNOMAD database for each ancestry^35^.

#### Genetic ancestry analysis

We used the UKBB genotype array data to perform principal component analysis (PCA). We first pruned the genotype data using the plink command ‘--indep-pairwise 500 50 0.05’. We then used FlashPCA^36^ based on 35,091 pruned variants. We merged the UKBB samples with 2,504 participants of 1000 Genomes Project (1KG phase 3)^37^ and kept only shared variants between the two datasets. Then, we used a random forest machine learning based on 10 principal components to train ancestry classifiers using 1KG labeled data. Finally, we then used the trained model to predict the genetic ancestry of the UKBB samples (**Supplementary Figure 1 (a, b)**).

### All of Us (AoU)

The AoU research program launched recruitment in 2018 across 340 sites across the United States and over 372,380 participants were enrolled by 2022. AoU combines participant-derived data from surveys such as self-reported health information, physical measurements, electronic health records, and biospecimens. We analyzed the AoU data on Workbench, a cloud-based environment^38^. The first data release included N=98,622 participants with complete SNP microarray and genome sequencing data as well as phenotype information. The participants included 60% female, the mean age was 55 years and consisted of 53% European, 4% Asian, and 21% Black/African American race by self-report. In addition, 17% of the cohort self-reported having Hispanic/Latinx ethnicity (**Table 1**).

#### SNP Microarray genotype data

All participants were genotyped with the Illumina Global Diversity Array (GDA). This microarray contains 1,904,679 SNVs and 44,172 indels. First, we performed genome-wide imputation analysis on the Workbench platform. Before imputation, we excluded all variants with MAF ≤ 0.005 (671,685 variants) or genotype missingness rate ≥ 0.05 (41,526 variants). We successfully lifted over genomic positions from human GRCh38 (hg38) to GRCh19 (hg19) for 96% of SNPs. We then adopted the TopMed pre-imputation quality control (QC) pipeline to correct allele designations and additionally remove poorly mapping variants^39^. After QC, we used 1,191,468 variants for imputation. To reduce RAM usage and increase speed, we split the 165,208 subjects with microarray data into 8 equal batches and then imputed each batch separately. After pre-phasing with EAGLE v.2^40^, we imputed missing genotypes using Minimac4^31^ and 1KG phase 3v5^41^ reference panel. A total of 43,371,225 autosomal variants were imputed in 165,208 individuals (**Supplementary Table 1)**. We then merged the eight batches based on position using VCFtools software with the command ‘vcftools --gzvcf --positions --recode --recode-INFO-all –stdout’. MAFs for the imputed markers were closely correlated (correlation coefficient (r) = 0.96) with the MAFs for the 1KG dataset.

#### Genetic ancestry analysis

Similar to the UKBB data, we first pruned the genetic data using the command ‘--indep-pairwise 500 50 0.05’ in PLINK^42^ and used N=36,358 pruned variants for kinship and ancestry analysis. Using KING software^43^, we removed 270 samples with pairwise kinship coefficients>0.35. We then merged our AoU samples with 1KG samples, kept only SNPs in common between the two datasets, calculated PCs for the 1KG samples, and projected each of our samples onto those PCs. We then used a random forest-based machine learning approach to assign a continental ancestry group to each AoU sample. Briefly, we trained and tested the random forest algorithm on 1KG subjects with known labels. We trained the random forest model using 10 PCs as a labeled feature matrix. Then we used our trained random forest model to predict the genetic ancestries for the AoU dataset (**Supplementary Table 2 and Supplementary Figure 1 (c, d)**).

#### Whole genome sequencing

We utilized 98,622 whole genome sequencing (GS) data released on March 15, 2020. The detailed description of GS is available elsewhere^44^. Briefly, the GS data were generated with NovaSeq 6000. DRAGEN v3.4.12 (Illumina) was used for genome alignment and calling, providing 702,668,125 SNVs for 98,622 samples with mean coverage greater or equal to 30x and >90% of bases at 20x coverage. The GS data is available in the All of Us workbench in the Hail matrix. We extracted all variants in *PKD1, PKD2*, *COL4A3*, *COL4A4*, and *COL4A5* genes in VCF format using the following hail command in Jupyter Notebook:

‘Gene_intervals = [’chr16:2.10M-2.15M’, ’chr4:87M-89M’,’chr2:220M-235M’,’chrX:107M-109M’] mt = hl.filter_intervals(

mt, [hl.parse_locus_interval(x,)
for x in Gene_intervals])

hl.export_vcf(mt, output_location, tabix=True)’

We then converted the vcf format data to the bed/bim/fam format using PLINK software^42^.

### Rare variant quality control, filtering, and classification

We analyzed genetic variants in protein-coding regions of two ADPKD genes (*PKD1* and *PKD2*) and three COL4A-AN genes (*COL4A3*, *COL4A3*, and *COL4A5*) in the UKBB and AoU datasets. We first removed variants with low genotype quality (GQ<90), depth of coverage (DP<10), and synonymous variants. Next, we filtered variants based on frequency in any ancestral groups across the UKBB, AoU, and gnomAD datasets^45^, excluding variants with MAF>0.00001 for *PKD1* and *PKD2* (considering the autosomal dominant inheritance of ADPKD) and MAF>0.001 for *COL4A3, COL4A4,* and *COL4A5* (considering the recessive inheritance of the most severe COL4A-AN phenotypes). We next used a range of prediction scores to define qualifying variants (QV), as recently proposed^46^. First, we identified all rare predicted loss of function (pLOF) variants, including stop-gain, frameshift, stop-lost, start-lost, and essential splice variants as defined previously^46^. Second, we classified rare missense variants as deleterious if they met the following strict criteria: 1) Revel score >0.70^47^ and 2) variants predicted as damaging by the consensus of five predictors: Sorting Intolerant from Tolerant (SIFT)^48^, Polymorphism Phenotyping v2 (PolyPhen2) HDIV and PolyPhen2 HVAR^49^; likelihood ratio test (LRT)^50^; and MutationTaster^51^. After defining the lists of pLOFs and predicted deleterious missense variants, we intersected these variants with ClinVar and Varsome databases and excluded all variants previously reported as ‘Benign’ (B) or ‘Likely Benign’ (LB) by at least one of these databases^52, 53^. Third, we identified all additional rare variants reported as ‘Pathogenic’ (P) or ‘Likely Pathogenic’ (LP) by at least two independent ClinVar submitters (accessed on 11/13/22). To increase the specificity, we excluded any variants with a conflict of reported pathogenicity or those submitted to ClinVar by only a single submitter. Based on these annotations, we then analyzed the data defining carrier status by three distinct variant classification models: the most stringent model (M1) included only pLOF and reported ‘P’ variants as defined above; model 2 (M2) was relaxed also to include pLOF, ‘P’, and ‘LP’ variants; and model 3 (M3) was further relaxed to include pLOF, and all deleterious missense variants predicted as deleterious by all 5 algorithms, with revel score > 0.7, and not previously classified as ‘B’ or ‘LB’ by ClinVar. For COL4A-AN analyses, we additionally analyzed a biallelic (recessive) inheritance by defining homozygous or compound heterozygous (*COL4A3* and *COL4A4)* or hemizygous (for *COL4A5* in males) genotypes for the qualifying variants. The list of observed qualifying variants included under each model is provided in **Supplemental Data 1 and 2.**

### Genome-wide Polygenic Score (GPS)

We used our validated GPS for CKD that was previously optimized for trans-ancestry performance^29^. The score was calculated using the PLINK command ‘--bfile --score sum --out’ based on imputed genotype data. The GPS distribution was ancestry-adjusted for mean and variance based on 1KG reference, normal standardized, and additionally adjusted for *APOL1* risk genotype as previously proposed (**Supplementary Figure 2**)^29^. The *APOL1* risk alleles were imputed for all subjects, and the risk genotype was defined under a recessive model as G1G1, G2G2, or G1G2 risk allele combinations across all datasets (**Supplementary Table 3**).

### CKD phenotyping and case-control definitions

We used our validated CKD e-phenotyping algorithm to define CKD cases and controls^54^. All cases had either estimated glomerular filtration rate (eGFR) below 60 ml/min/1.73 m^2^ (by 2009 CKD-EPI equation^55^) or received renal replacement therapy (dialysis or kidney transplant). All controls had eGFR greater than 90 ml/min/1.73 m^2^ and no evidence of CKD based on diagnostic or procedure billing codes. Additional covariates included age, sex, diabetes (type I or type II), and significant principal components of ancestry, similar to our published GPS validation studies^29^.

### Predictive performance

The predictive performance of the GPS was assessed using standardized metrics as recently proposed by ClinGen^56^, including area under the receiver operating characteristics curve (AUROC), variance explained (R^2^), and effect size (OR) per standard deviation of the GPS distribution in controls. We used the pROC R package to calculate AUROC. For effect size estimation, we used logistic regression (glm function in R) with CKD status as an outcome and standardized GPS as a predictor with adjustment for age, sex, diabetes mellitus (type I or type II), genotype/imputation batch, and four PCs of ancestry, similar to prior studies^29^. Similarly, the association of a carrier status with CKD was tested using a logistic regression with CKD case status as an outcome and carrier status as a predictor, controlling for age, sex, diabetes, batch, and ancestry PCs. The same logistic model with the included GPS and carrier status terms was used to test the GPS-by-carrier status interaction. To compare GPS effect sizes between carriers and non-carriers, we derived ORs (and 95% CIs) of CKD, comparing each tertile of the GPS distribution in the carriers to the reference middle (2^nd^) tertile of the GPS for noncarriers in each cohort. For all analyses, we used R version 4.2.2 (2022-10-31).

### Meta-PheWAS

We performed a phenome-wide association analysis for ADPKD and COL4A-AN variant carriers in both AoU and UKBB datasets. The 165,208 genotyped and imputed AoU participants had 12,945 International Classification of Diseases, Ninth Revision, Clinical Modification (ICD-9-CM) codes that were first mapped to 1,817 distinct phecodes. Similarly, there were 10,221 ICD-9 codes for UK Biobank participants (N=460,363) with imputed genotype data that mapped to 1,817 distinct phecodes. Phenome-wide associations were performed using the PheWAS R package^57^. The package uses two ICD-9 codes occurrences within a given phecode grouping to define a case and pre-defined “control” groups for each phecode. All 1,817 phecodes were tested using logistic regression with case-control status as the outcome and genotype, sex, age, batch, and five principal components of ancestry as predictors. We then performed fixed effects Meta-PheWAS of AoU and UKBB datasets using the PheWAS R package. We set the Bonferroni corrected statistical significance threshold for phenome-wide significance at 2.75E-05 (0.05/1,817 phecodes tested).

## RESULTS

The summary of our analytical approach is provided in **Figure 1** as a flowchart. Using our electronic phenotyping algorithms, we defined a total of 10,081 CKD cases and 266,724 controls in the UKBB and 11,820 CKD cases and 22,763 controls in the AoU dataset among those participants with both high-quality sequence and SNP microarray data available.

**Figure 1:**
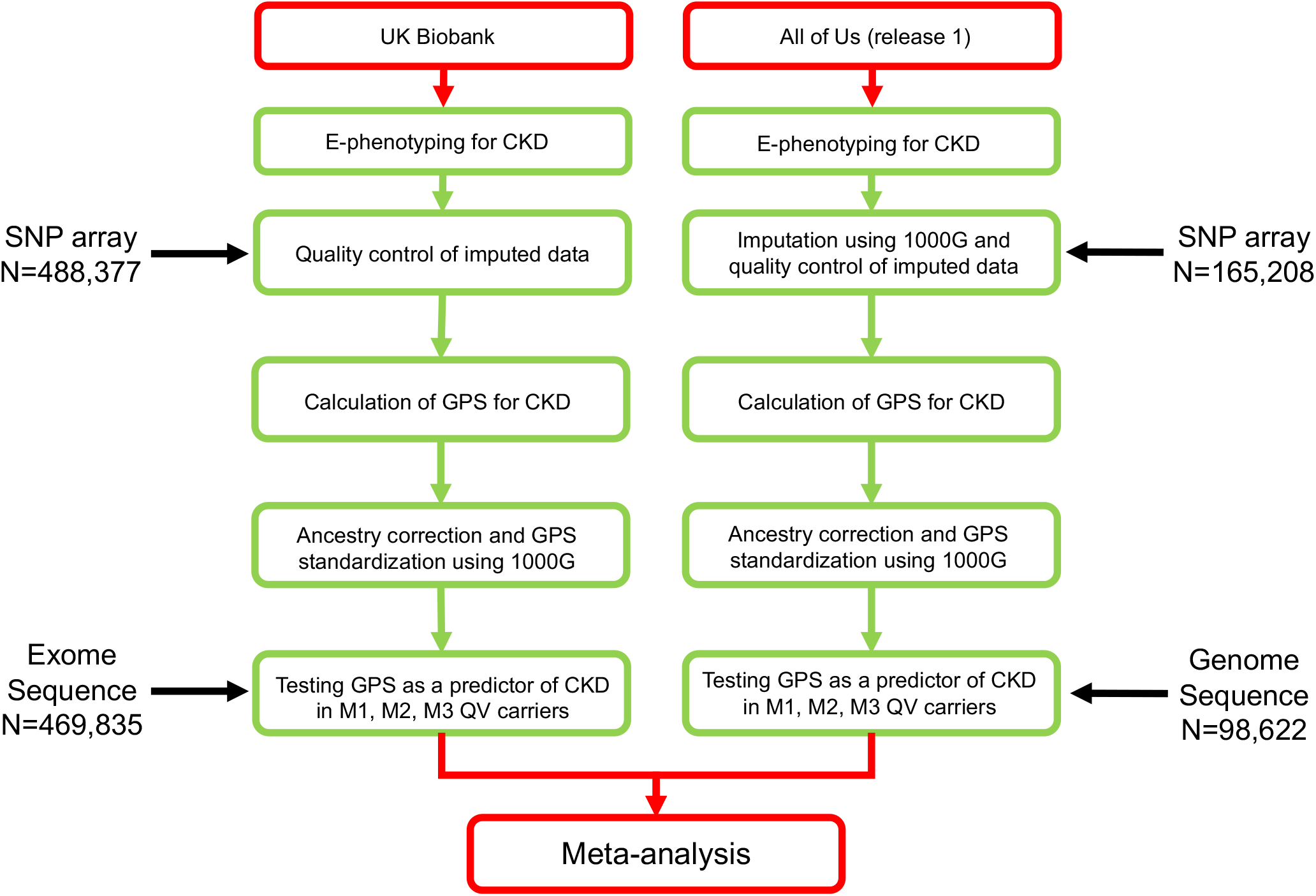
Overview of the workflow for the analysis of phenotype and genotype data.

### Autosomal Dominant Polycystic Kidney Disease (ADPKD)

We first identified all *PKD1* and *PKD2* variants that were either pLoF or reported as ‘P’ by at least two ClinVar submitters without conflicts (model M1). A total of 172 and 34 carriers of such variants were found in the UKBB and AoU, corresponding to the overall prevalence of approximately 0.036% and 0.034%, respectively. We performed a Meta-PheWAS analysis of both UKBB and AoU datasets to assess phenome-wide associations of M1 variants (**Figure 2a)**. The top associated phecode was “Cystic Kidney Disease” with OR=295.7 (95%CI: 214.3-408.0, P=9.0E-263), as expected. We also detected significant associations with a variety of CKD-related phecodes, including “End-stage renal disease”, OR=52.8 (95%CI: 31.2-89.3, P=2.1E-49) and “Kidney replaced by transplant” OR=112.1 (95%CI: 71.5-175.7, P=4.9E-94), as well as multiple other renal and extra-renal complications of ADPKD (**Supplemental Data 3)**, confirming that M1 variant definitions have robust phenotypic signatures across both biobanks. We also tested the effects of these variants on the risk of CKD, as defined by our phenotyping algorithm, after adjustment for age, sex, diabetes, batch, and ancestry (**Supplementary Table 4**). In the meta-analysis of both cohorts, the risk of CKD was 17-fold higher in the ADPKD M1 variant carriers compared to non-carriers (OR: 17.1, 95%CI: 11.1-26.4, P=1.8E-37).

**Figure 2:**
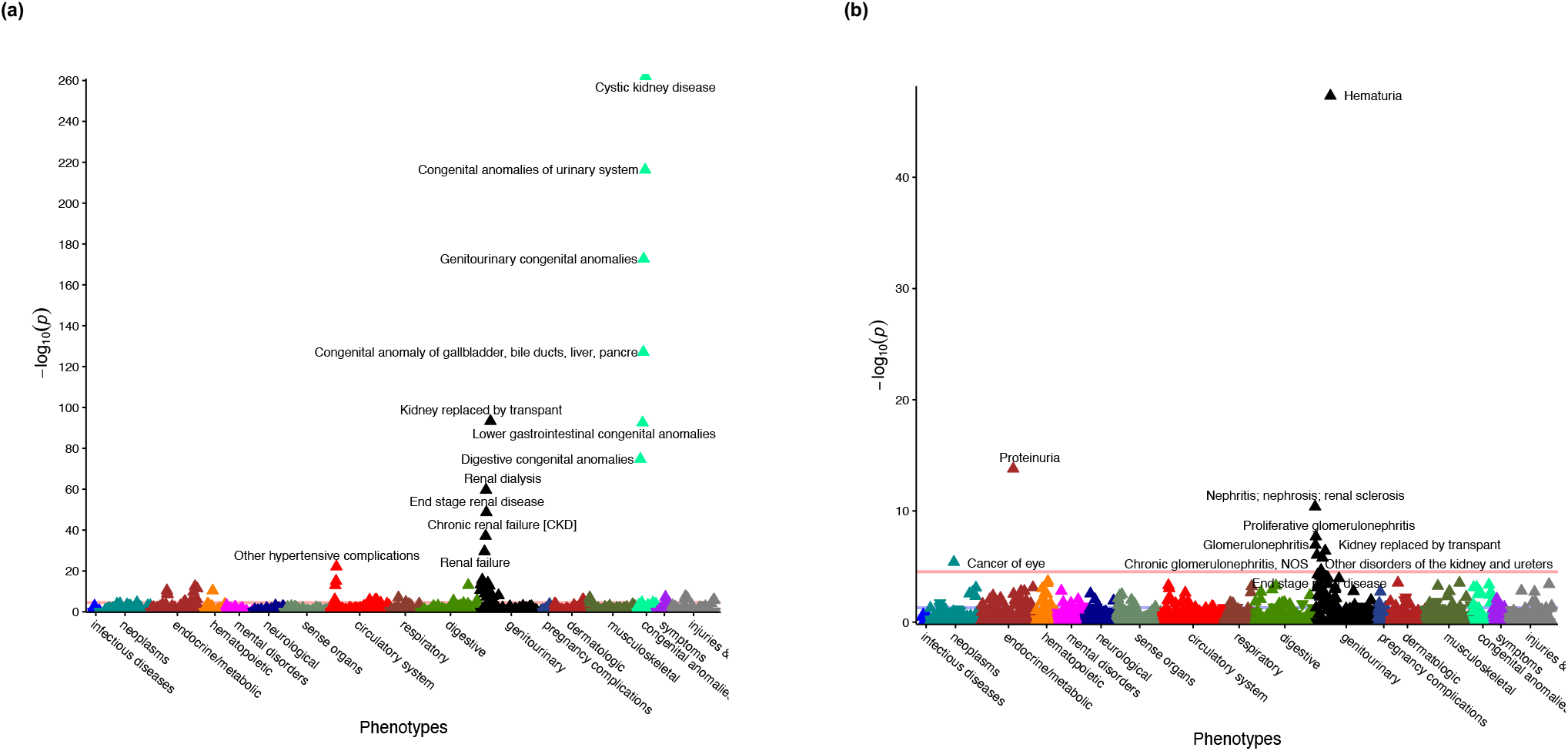
Meta-PheWAS for (a) ADPKD M1 variant carriers and (b) COL4A-AN M1 variant carriers. This meta-analysis includes combined data from 460,360 UKBB and 74,350 AoU participants, with both genotype and phenotype data available. Both analyses were performed under the dominant inheritance model and adjusted for age, sex, diabetes, batch, and ancestry. The red horizontal lines indicate a phenome-wide significance level after accounting for the number of phecodes tested (P=2.8E-05). Y-axis: -log10(P-value) from fixed effects meta-analysis. P-values are two-sided and provided without accounting for multiple testing. X-axis: system-based phecode groupings. An upward-pointing triangle indicates increased odds for a given phecode, downward-pointing triangle indicates reduced risk.

We next investigated the effect of polygenic background on the risk of CKD by computing our previously validated GPS for CKD^29^ in all UKBB and AoU participants. After *APOL1* and ancestry adjustments, the polygenic score was standard normal-distributed across ancestries in both UKBB and AoU datasets (**Supplementary Figure 2**). Because this risk score has not been previously tested in AoU participants, we first confirmed that the GPS was indeed associated with increased risk of CKD in this dataset (OR per SD=1.39, 95%CI: 1.36-1.43, P=5.9E-125, adjusted for age, sex, diabetes, batch, and genetic ancestry). All participants were then stratified based on their ADPKD QV carrier status, and the effects of the GPS were re-examined within each stratum across both UKBB and AoU datasets combined. In the meta-analysis, the OR per SD of the GPS was 2.28 (95%CI: 1.55-3.37, P=2.7E-05) in the M1 QV carriers compared to 1.72 (95%CI: 1.69-1.76, P<E-300) in the non-carriers (**Table 2**). Despite the trend for a greater effect of the GPS among the carriers, the GPS-by-carrier interaction test was not statistically significant in either cohort or in the combined meta-analysis.

**Table 2:** Performance metrics for the GPS in ADPKD M1, M2, and M3 carriers and non-carriers in the meta-analysis of UKBB and AoU cohorts. . OR adjusted for age, sex, diabetes, PCs of ancestry, and genotyping array or batches; AUC was calculated for the full model (GPS and covariates) and for GPS alone without covariates (crude); variance explained was calculated for the GPS alone by estimating variance explained by the full model (GPS and covariates) minus the variance explained by the covariates-only model. P-values are two-sided and not corrected for multiple testing. CI: Confidence Intervals.

| ADPKD variants | Cases/controls | OR (95% CI), P-value | AUC full model (95%CI) | AUC crude (95%CI) | Variance explained |
| --- | --- | --- | --- | --- | --- |
| **Non-carrier** |  |  |  |  |  |
|  | 21,901/275,638 | 1.72 (1.69-1.76), P<E-300 | 0.78 (0.78-0.78) | 0.62 (0.62-0.62) | 0.039 |
| **M1** |  |  |  |  |  |
|  | 41/81 | 2.28 (1.55-3.37), P=2.6E-05 | 0.96 (0.92-1.00) | 0.69 (0.59-0.79) | 0.128 |
| **M2** |  |  |  |  |  |
|  | 44/95 | 2.21 (1.37-3.58), P=3.3E-05 | 0.97 (0.93-1.00) | 0.70 (0.60-0.80) | 0.103 |
| **M3** |  |  |  |  |  |
|  | 52/215 | 5.25 (2.31-11.9), P=7.4E-05 | 0.97 (0.94-1.00) | 0.69 (0.60-0.78) | 0.076 |

We next estimated the CKD risk for each tertile of the GPS distribution among the M1 variant carriers compared to the middle tertile of the non-carriers (i.e., reflecting average risk) across both AoU and UKBB (**Figure 3** and **Supplementary Table 5**). Among the QV carriers, we observed a clear gradient of CKD risk as a function of GPS, ranging from OR=3.03 (95%CI 1.03-8.95, P=4.4E-02) for the lowest tertile to OR=54.4 (95%CI 26.1-113.0, P=9.6E-27) for the highest tertile of polygenic risk. These results demonstrate that the GPS significantly alters the penetrance of ADPKD M1 qualifying variants.

**Figure 3.**
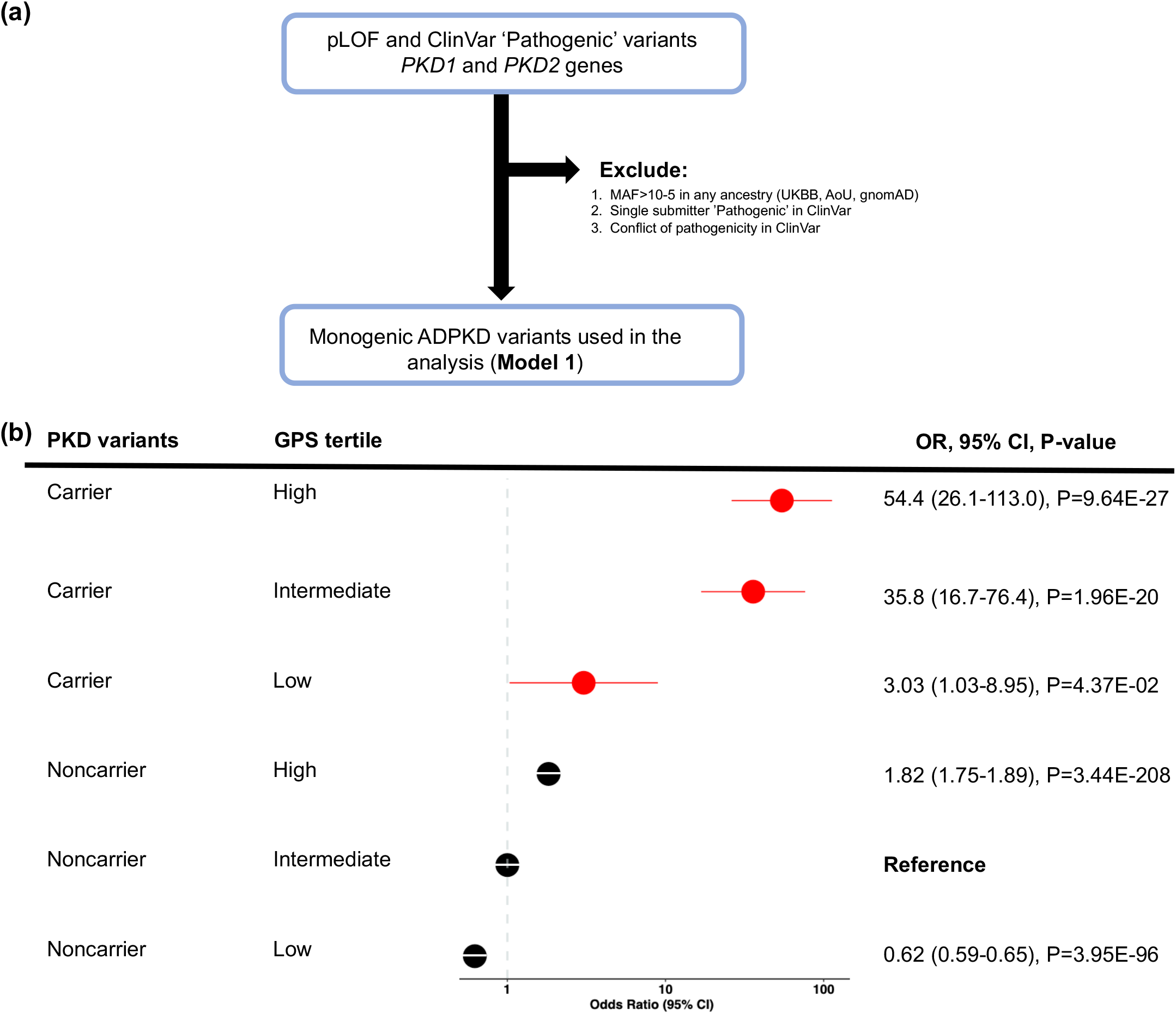
Polygenic effects on the risk of CKD among ADPKD M1 variant carriers (dominant model): **(a)** M1 qualifying variant filtering strategy; **(b)** CKD risk for each polygenic risk score tertile compared to the middle tertile of non-carriers (average population risk). The analysis includes N=262,435 UKBB participants (N_cases=_9,565 and N_controls=_252,870) and N=34,603 AoU participants (N_cases=_11,830 and N_controls=_22,773). The non-carriers with intermediate polygenic scores (middle tertile) served as the reference group for all calculations. X-axis shows Odds Ratios; the dotted vertical line corresponds to the OR=1.0 (no change in risk compared to the reference). P-values correspond to the fixed effects meta-analysis between UKBB and AoU cohorts. P-values are two-sided and are not corrected for multiple testing. GPS: Genome-wide Polygenic Score.

In the subgroup analyses, we examined QVs in *PKD1* and *PKD2* separately and observed similar patterns of GPS effects within each of the gene-defined subgroups (**Supplementary Figure 3**). Similarly, we examined QVs by variant type (truncating vs. missense) and observed a consistent pattern of GPS effects for both subgroups (**Supplementary Figure 4**). Lastly, we investigated the effect of the GPS on the risk of CKD among ADPKD carriers defined under alternative QV models (M2 and M3, **Supplementary Table 5**). Similar results on the penetrance of CKD were observed, demonstrating that our findings were also robust to less stringent QV definitions.

### Collagen IV Alpha Associated Nephropathy (COL4A-AN)

We next examined the effect of GPS on the risk of CKD in the carriers of COL4A-AN variants compared to the average risk of non-carriers. In this analysis, we used a less stringent MAF<0.001 for variant filtering, considering that the most severe phenotype of COL4A-AN is observed under a recessive model. Under M1, we defined a total of 1,435 carriers in the UKBB and 310 carriers in the AoU dataset, corresponding to the overall prevalence of approximately 0.31% and 0.32%, respectively.

In the Meta-PheWAS analysis for M1 carriers across both UKBB and AoU datasets (**Figure 2 (b))**, the top associated phecode was “Hematuria” with OR=2.3 (95% CI: 2.0-9.6, P=4.8E-48). Other phenome-wide-significant associations included “Kidney replaced by transplant” (OR=3.1, 95%CI: 2.0-23.8, P=3.8E-07), “Nephritis, nephrosis, renal sclerosis” (OR=2.34, 95%CI: 1.81-10.39, P=4.1E-11), “Proteinuria” (OR=3.94, 95%CI: 2.77-51.6, P=1.6E-14) and “Chronic glomerulonephritis, NOS” (OR=2.98, 95%CI: 1.92-19.7, P=9.0E-07). The complete list of phenotypic associations is provided as **Supplemental Data 4**. Compared to non-carriers, the M1 QV carriers had a 37% increased risk of CKD as defined by our e-phenotype (OR=1.37, 95%CI: 1.13-1.64, P=8.5E-04), M2 carriers had 25% increased risk (OR=1.25, 95%CI: 1.00-1.56, P=4.9E-02), and M3 carriers had 48% increased risk (OR=1.48, 95%CI: 1.23-1.77, P=2.6E-05) in the combined meta-analysis under a dominant model (**Supplementary Table 6).** In comparison, the M3 recessive genotype was associated with a 3.38-fold higher risk (OR=3.38, 95%CI: 1.88-6.08, P=4.7E-05).

We next investigated the effect of polygenic background on the risk of CKD among M1 QV carriers compared to noncarriers. Similar to ADPKD, the GPS had a significant effect on the risk of CKD among both COL4A-AN carriers (OR per SD of GPS = 1.78, 95%CI: 1.22-2.58, P=2.4E-03) and non-carriers (OR per SD of GPS =1.70, 95%CI: 1.68-1.73, P<E-300) in the meta-analysis (**Table 3**). There was no significant GPS-by-carrier interaction (P=8.1E-01). Similar to ADPKD, we observed a gradient of CKD risk as a function of the GPS among M1 carriers, from no increased risk (OR=1.01, 95%CI 0.63-1.86, P=7.8E-01) for the lowest GPS tertile to a 2.5-fold higher risk (OR=2.53, 95%CI 1.66-3.85, P=1.4E-05) for the top GPS tertile when compared to the middle tertile of non-carriers (**Figure 4**).

**Table 3:** Performance metrics for the GPS in COL4A-AN M1, M2, and M3 carriers and non-carriers in the meta-analysis of UKBB and AoU; OR adjusted for age, sex, diabetes, PCs of ancestry, and genotyping array or batches; AUC was calculated for the full model (GPS and covariates) and for GPS alone without covariates (crude); variance explained was calculated for the GPS alone by estimating variance explained by the full model (GPS and covariates) minus the variance explained by the covariates-only model. P-values are two-sided and not corrected for multiple testing. CI: Confidence Intervals.

| COL4A-AN variants | Cases/controls | OR (95% CI), P-value | AUC full model (95%CI) | AUC crude (95%CI) | Variance explained |
| --- | --- | --- | --- | --- | --- |
| **Non-carrier** |  |  |  |  |  |
|  | 21,901/275,638 | 1.70 (1.68-1.73), P<E-300 | 0.77 (0.77-0.77) | 0.63 (0.63-0.63) | 0.038 |
| **M1** |  |  |  |  |  |
|  | 99/1193 | 1.78 (1.22-2.58), P=2.4E-03 | 0.94 (0.91-0.97) | 0.59 (0.52-0.65) | 0.019 |
| **M2** |  |  |  |  |  |
|  | 112/1,344 | 2.47 (1.56-3.94), P=1.3E-04 | 0.93 (0.90-0.96) | 0.62 (0.56-0.68) | 0.014 |
| **M3** |  |  |  |  |  |
|  | 172/1,884 | 1.93 (1.26-2.95), P=2.3E-03 | 0.89 (0.86-0.92) | 0.60 (0.55-0.65) | 0.019 |

**Figure 4.**
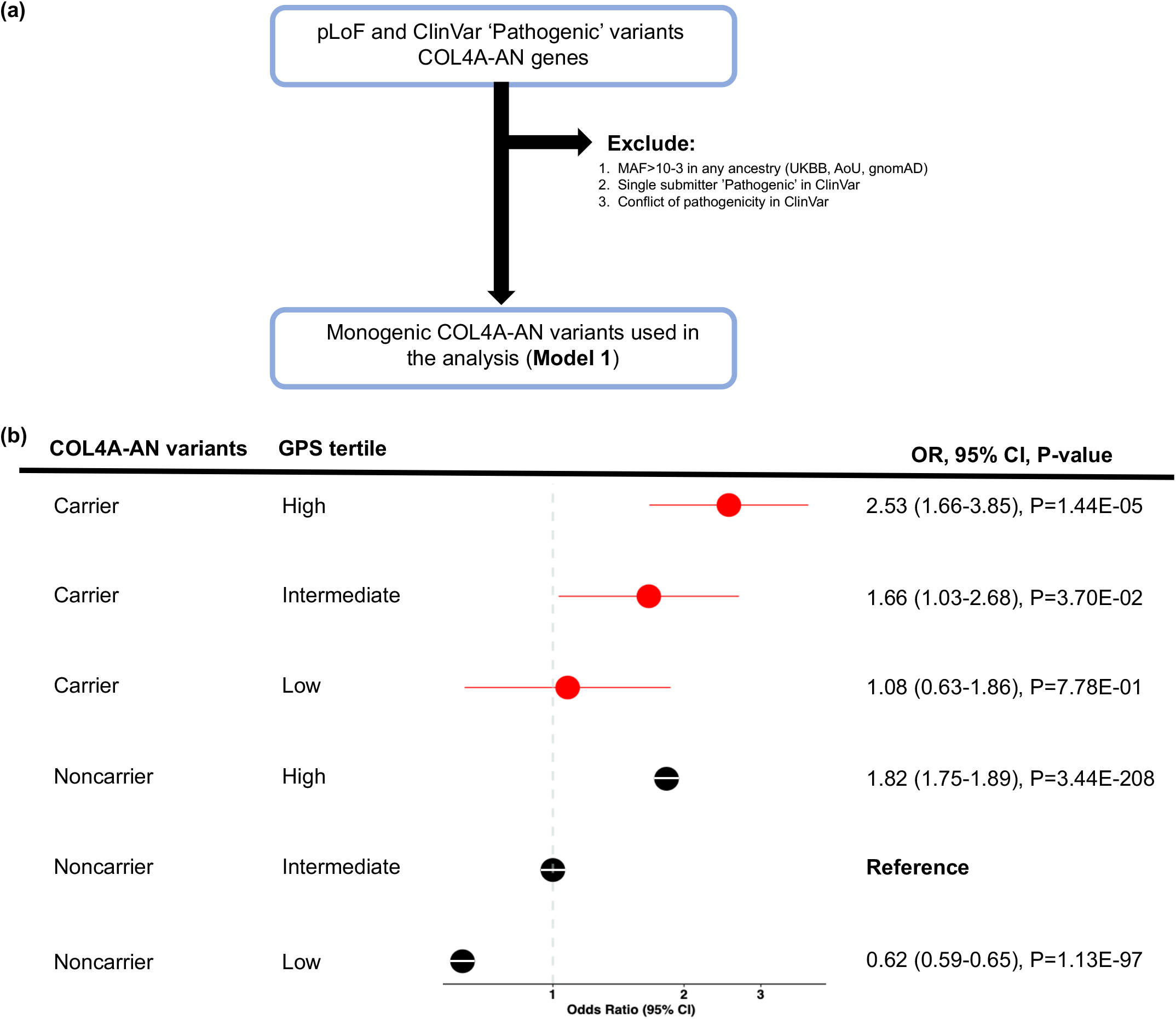
Polygenic effects on the risk of CKD among M1 carriers of COL4A-AN variants (dominant model): **(a)** M1 qualifying variant filtering strategy; **(b)** CKD risk for each polygenic score tertile compared to the middle tertile in non-carriers (average population risk). The analysis includes N=262,435 UKBB participants (N_cases=_9,565 and N_controls=_252,870) and N=34,603 AoU participants (N_cases=_11,830 and N_controls=_22,773). The non-carriers with intermediate polygenic risk (middle tertile) served as the reference group for all calculations. X-axis shows odds ratios; the dotted vertical line corresponds to the OR=1.0 (no change in risk compared to the reference). P-values correspond to the fixed effects meta-analysis between the two cohorts. P-values are two-sided and are not corrected for multiple testing. GPS: Genome-wide Polygenic Score.

We also explored the recessive model by testing for GPS effects among individuals with the M3 risk genotype (QV homozygotes, compound heterozygous, or *COL4A5* hemizygous males). For individuals with the risk genotype, the top tertile of the GPS conveyed a 6.73-fold higher risk of CKD (OR=6.73, 95%CI: 2.59-17.5, P=8.8E-05), while the bottom tertile conveyed a 2.29-fold higher risk of CKD (OR=2.29, 95%CI 0.64-8.12, P=2.0E-01) compared to the middle tertile of individuals without the risk genotype (**Figure 5**).

**Figure 5.**
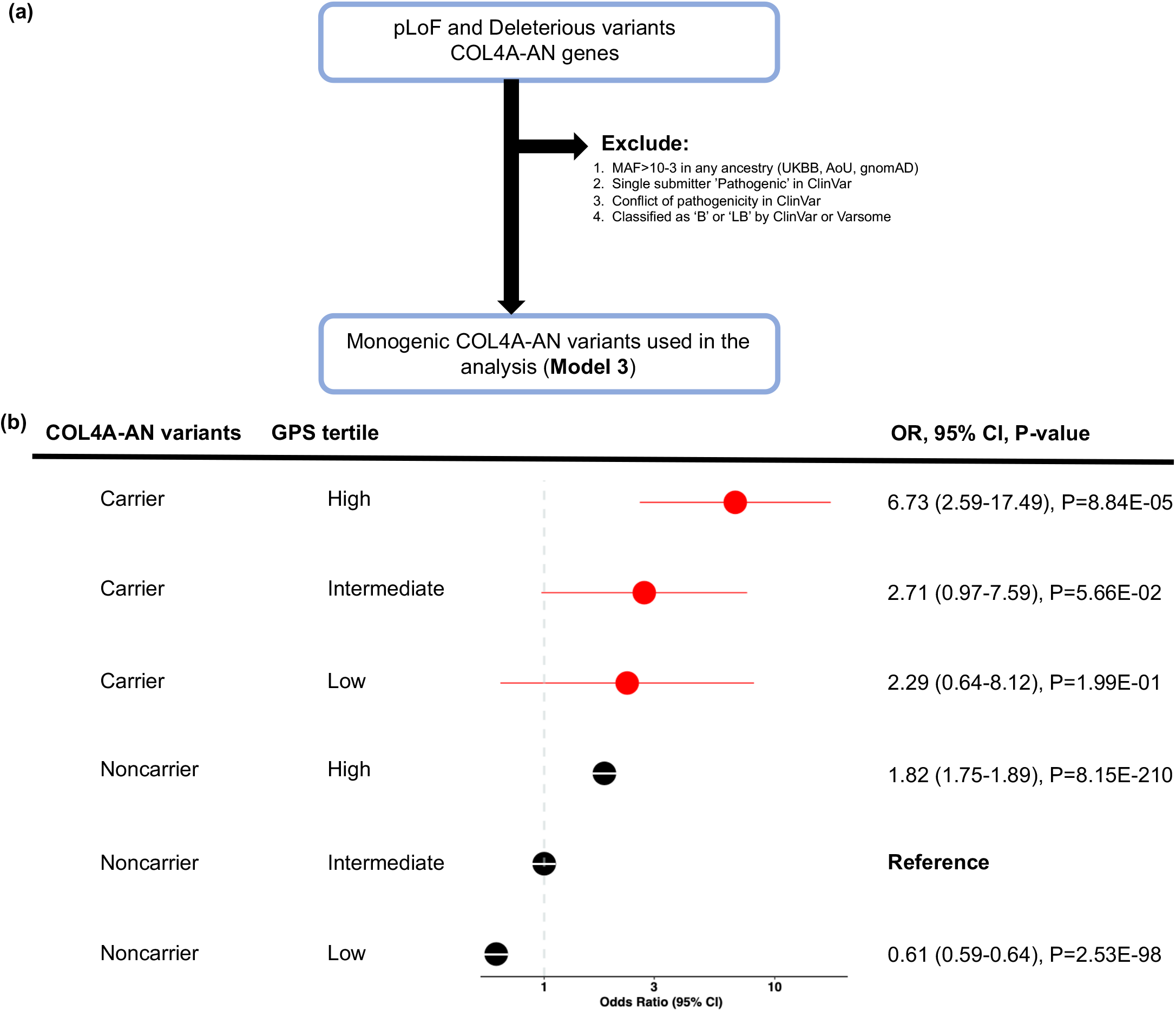
Polygenic effects on the risk of CKD among biallelic carriers of COL4A-AN M3 variants (recessive model): **(a)** M3 qualifying variant filtering strategy; **(b)** CKD risk for each polygenic risk score tertile compared to the middle tertile in non-carriers (average population risk). The analysis includes N=262,435 UKBB participants (N_cases=_9,565 and N_controls=_252,870) and N=34,603 AoU participants (N_cases=_11,830 and N_controls=_22,773). The non-carriers with intermediate polygenic score (middle tertile) served as the reference group for all calculations. X-axis shows odds ratios; the dotted vertical line corresponds to the OR=1.0 (no change in risk compared to the reference). P-values correspond to the fixed effects meta-analysis between the two cohorts. P-values are two-sided and are not corrected for multiple testing. GPS: Genome-wide Polygenic Score.

Our sensitivity analyses included alternative variant models (**Supplementary Table 7**) and individual analyses of autosomal (*COL4A3* and *COL4A4*) and sex-linked (*COL4A5*) genes (**Supplementary Table 8).** These analyses confirmed the direction-consistent effect of the GPS across all different subgroups. We note that recessive analyses for M1 and M2 models were underpowered due to the low overall frequency of recessive genotypes defined under these models.

## DISCUSSION

Our large-scale analyses of UKBB and AoU datasets demonstrated that polygenic background affects the risk of kidney disease among individuals with the most common forms of monogenic kidney disorders. This effect was most pronounced in the individuals with known pathogenic or rare pLOF variants in *PKD1* or *PKD2.* The bottom tertile of the GPS was associated with a 3-fold increased risk among these individuals compared to the middle tertile of non-carriers (average risk), while the top tertile was associated with a 54-fold increased risk of CKD. Similar but less extreme patterns of polygenic effects were also observed for COL4A-AN. The carriers of known pathogenic or rare pLOF variants in COL4A-AN genes in the bottom tertile of the GPS had no increased risk of CKD, while the individuals in the top GPS tertile had a 3-fold higher risk of CKD compared to non-carriers. Under the recessive model, the risk was 2-fold higher and nearly 6-fold higher for the bottom and top tertile of the GPS, respectively, compared to the average risk of non-carriers.

While our analyses suggest that the GPS significantly affects the penetrance of ADPKD and COL4A-AN, we recognize that our study has limitations. First, significant demographic differences exist between the UKBB and the AoU participants. The UKBB participants are older (mean age 56.5 years, range 40-69 years) and predominantly (94%) of European ancestry, while the AoU participants are younger (mean age 54.9 years, range 18-89 years) and have more diverse ancestral backgrounds (57% non-European). Because the risk of CKD increases with age, these differences may be partially responsible for a lower effect estimate for the GPS in the AoU compared to the UKBB dataset. Moreover, current catalogues of “P” and “LP” variants are more comprehensive for European compared to non-European populations. Thus, we are also more likely to misclassify pathogenic variants in the AoU dataset compared to the UKBB dataset, and such misclassification could also reduce the observed effect sizes.

Second, we were able to investigate only the two most common forms of monogenic kidney diseases, ADPKD and COL4A-AN. Similar patterns of GPS effects observed in these very different disorders suggest that our findings may be generalizable to other less frequent monogenic kidney diseases. However, much larger datasets would be needed to validate this hypothesis.

Third, we are aggregating qualifying variants across all known genes for ADPKD or COL4-AN. However, the penetrance of kidney disease is known to vary according to a specific gene (e.g., *PKD2* vs. *PKD1*) or a specific mutation type (e.g., missense vs. truncating variants). We performed sensitivity analyses to address this issue and our analyses by gene and variant type demonstrated consistent patterns of GPS effects across all subgroups. At the same time, we note that some of our subgroup analyses were underpowered. For example, *PKD2* mutations account for only ∼15-20% of ADPKD cases and lead to a less severe disease compared to *PKD1*^58^, impacting our power for individual analysis of this gene. Similarly, we do not have adequate power to define GPS effects under recessive inheritance using our most stringent (M1 and M2) models in COL4-AN. Thus, our biallelic analysis was limited to the M3 model.

Fourth, there are notable limitations when it comes to kidney disease phenotyping in large biobanks that relate to ascertainment biases, the cross-sectional nature of data, the non-random missingness of EHR diagnoses, and the inability to perform manual chart reviews to confirm the diagnosis. These and other limitations of our e-phenotyping strategy have been discussed elsewhere^54^. At the same time, the notable strength of our phenotyping approach is the fact that we are able to combine structured billing code data with all of the available laboratory tests to not only identify CKD cases but also to stage kidney disease severity with a high degree of confidence. Lastly, we recognize several important limitations of the GPS for CKD that was used here as a proxy for polygenic effects. These limitations have previously been described in depth elsewhere^29^. The effects of monogenic kidney disease demonstrated here will need to be re-assessed once more powerful polygenic scores for CKD become available.

In summary, in our combined analysis of SNP microarray, exome/genome sequencing, and EHR data, we observed significant additive effects of monogenic and polygenic factors on the risk of kidney disease across two large-scale biobanks. We demonstrated that in both ADPKD and COL4-AN, the risk of CKD could be either attenuated or amplified by the polygenic profile. We conclude that polygenic risk scores could be potentially used to improve our current risk stratification of patients with ADPKD and COL4-AN. Testing these findings in other forms of inherited kidney disorders will require further studies.

## Data Availability

The UKBB genotype and phenotype data are available through the UKBB web portal at https://www.ukbiobank.ac.uk/. The AoU genotype, WGS, and phenotype data are available through the AoU researcher workbench at https://www.researchallofus.org/data-tools/workbench/.

## Acknowledgments

This work was funded by the National Human Genome Research Institute (NHGRI) Electronic Medical Records and Genomics-IV (eMERGE-IV grant 5U01HG008680-07), National Library of Medicine (NLM grant R01LM013061), National Institute of Diabetes, Digestive and Kidney Diseases (NIDDK grant 5K25DK128563-03) and National Center for Advancing Translational Sciences (NCATS grant UL1TR001873). We are grateful to all participants from the UKBB and AoU projects for contributing their data and biological samples that enabled this study. The research on UK Biobank data has been conducted using the UK Biobank Resource under Application Number 41849. The All of Us Research Program is supported by the NIH Office of the Director through the following grants: Regional Medical Centers: 1OT2OD026549; 1OT2OD026554; 1OT2OD026557; 1OT2OD026556; 1OT2OD026550; 1OT2OD 026552; 1OT2OD026553; 1OT2OD026548; 1OT2OD026551; 1OT2OD026555; IAA# AOD16037; Federally Qualified Health Centers: HHSN 263201600085U; Data and Research Center: 5U2COD023196; Biobank: 1U24OD023121; The Participant Center: U24OD023176; Participant Technology Systems Center: 1U24OD023163; Communications and Engagement: 3OT2OD023205; 3OT2OD023206; and Community Partners: 1OT2OD025277; 3OT2OD025315; 1OT2OD025337; 1OT2OD025276.

## Author Contributions Statement

Project conceptualization: A.K., K.K.; analytical methods and genetic data analysis: A.K., K.K.; electronic phenotyping: N.S., C.W., G.H., J.G.N., monogenic variant filtering methods and review: A.G., P.C.H.; manuscript draft: A.K., K.K; overall project supervision: K.K.

## Code availability

The CKD phenotype software is available from the Phenotype Knowledge Database at https://phekb.org/phenotype/chronic-kidney-disease. The CKD GPS score equation is available through the PGS catalog at https://www.pgscatalog.org/publication/PGP000269/ and the Kiryluk Lab website: http://www.columbiamedicine.org/divisions/kiryluk/study_GPS_CKD.php.

## Supplementary Information

### Contents

#### Supplementary Figures

**Supplementary Figure 1.**
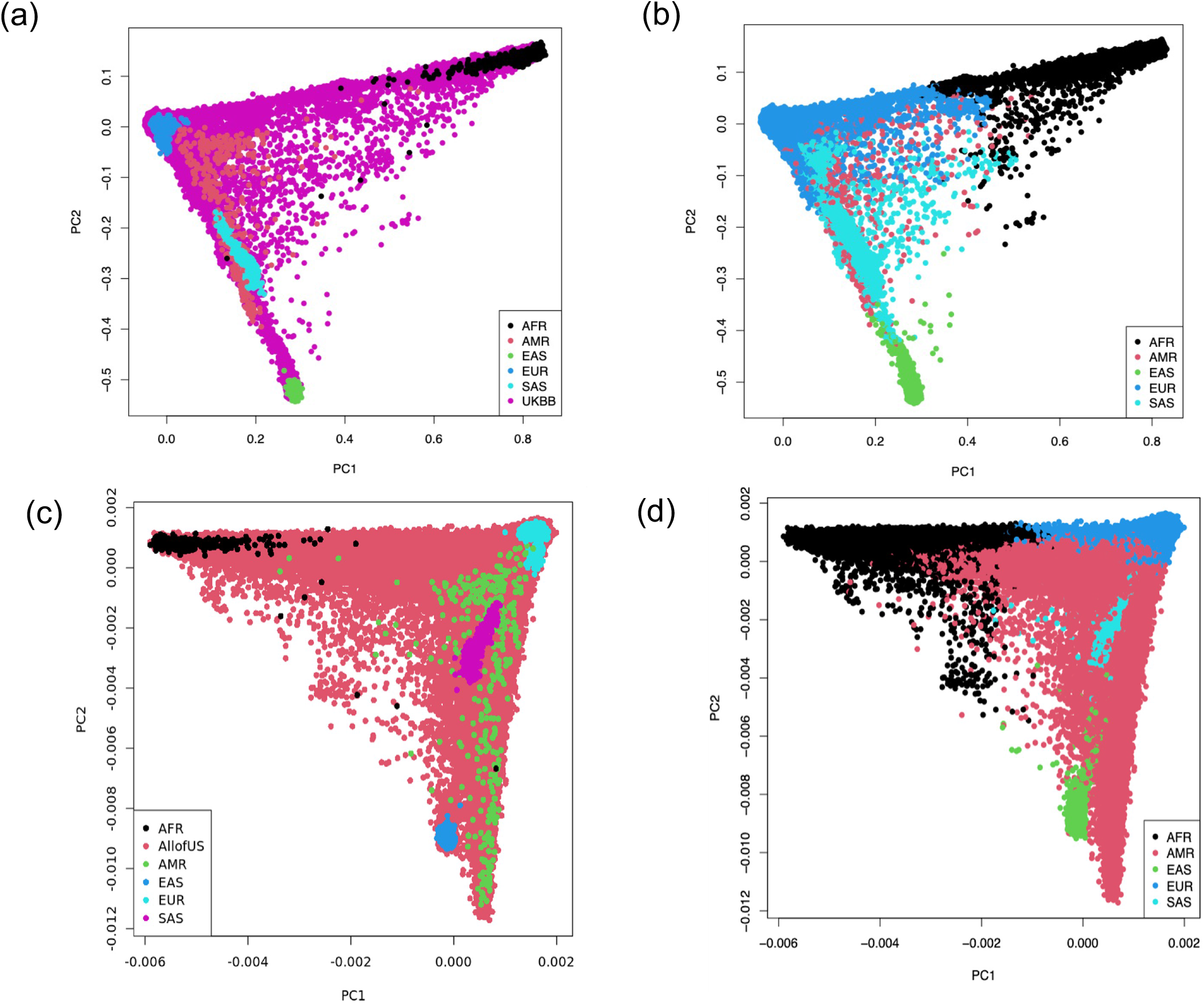
PCA projections of the study participants from the UKBB (top) and AoU (bottom) against the 1000 G reference populations: (a) UKBB (N = 460,360) and (c) AoU (N = 165,208) participants plotted against the reference 1000 G populations (N = 2,504), (b) machine learning-assigned ancestry for the UKBB and (d) the AoU datasets. X-axis: PC1; Y-axis: PC2; AFR: African; AMR: Admixed American; EAS: East Asian; EUR: European; and SAS: South Asian.

**Supplementary Figure 2.**
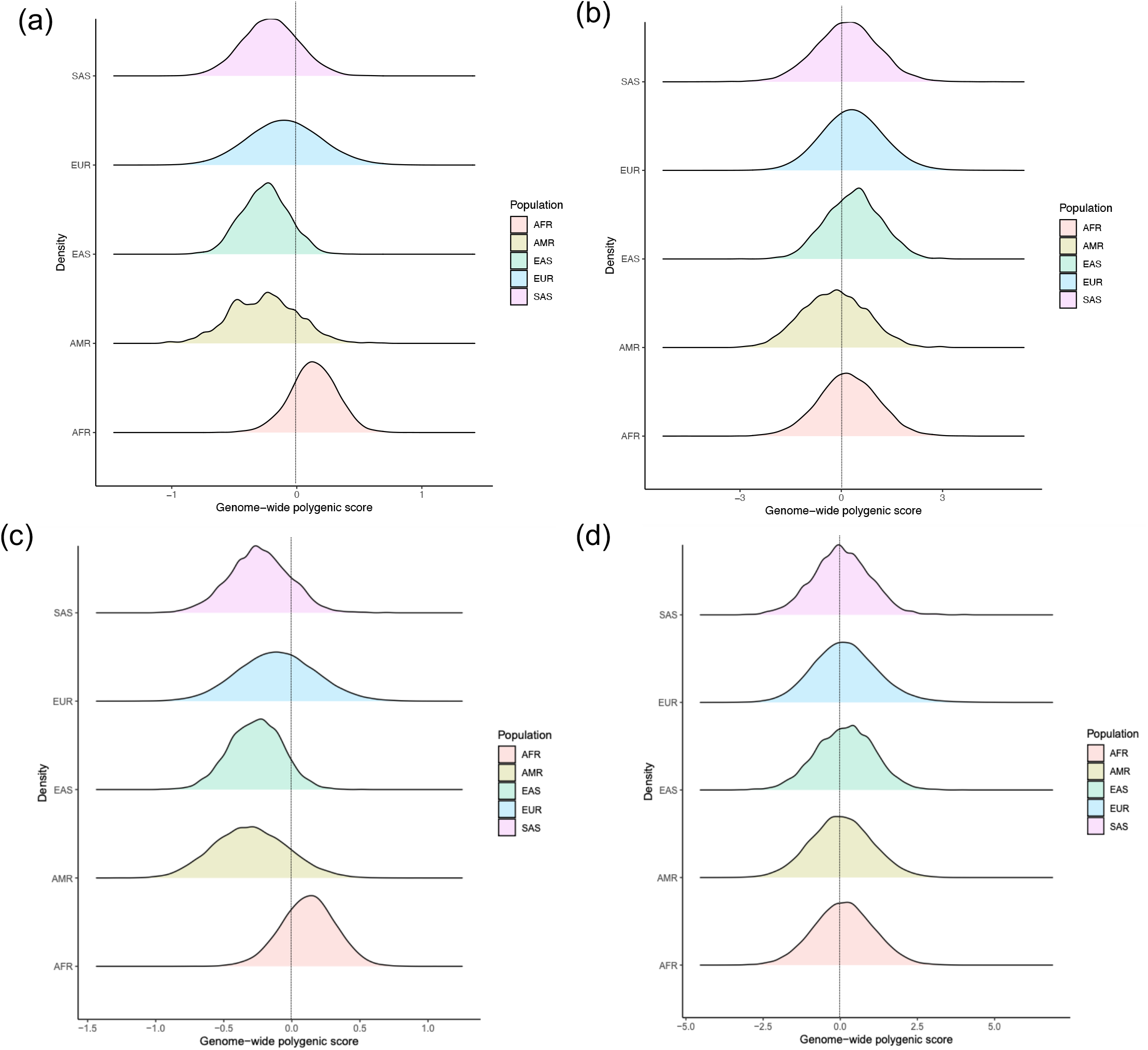
Genome-wide Polygenic Score (GPS) distributions by ancestry in the UKBB and AoU datasets: **(a)** unadjusted and **(b)** ancestry-adjusted GPS in UKBB (N = 460,360); **(c)** unadjusted and **(d)** ancestry-adjusted GPS in AoU (N = 165,208). EUR: European, AFR: African, AMR: Admixed American, EAS: East Asian, and SAS: South Asian.

**Supplementary Figure 3.**
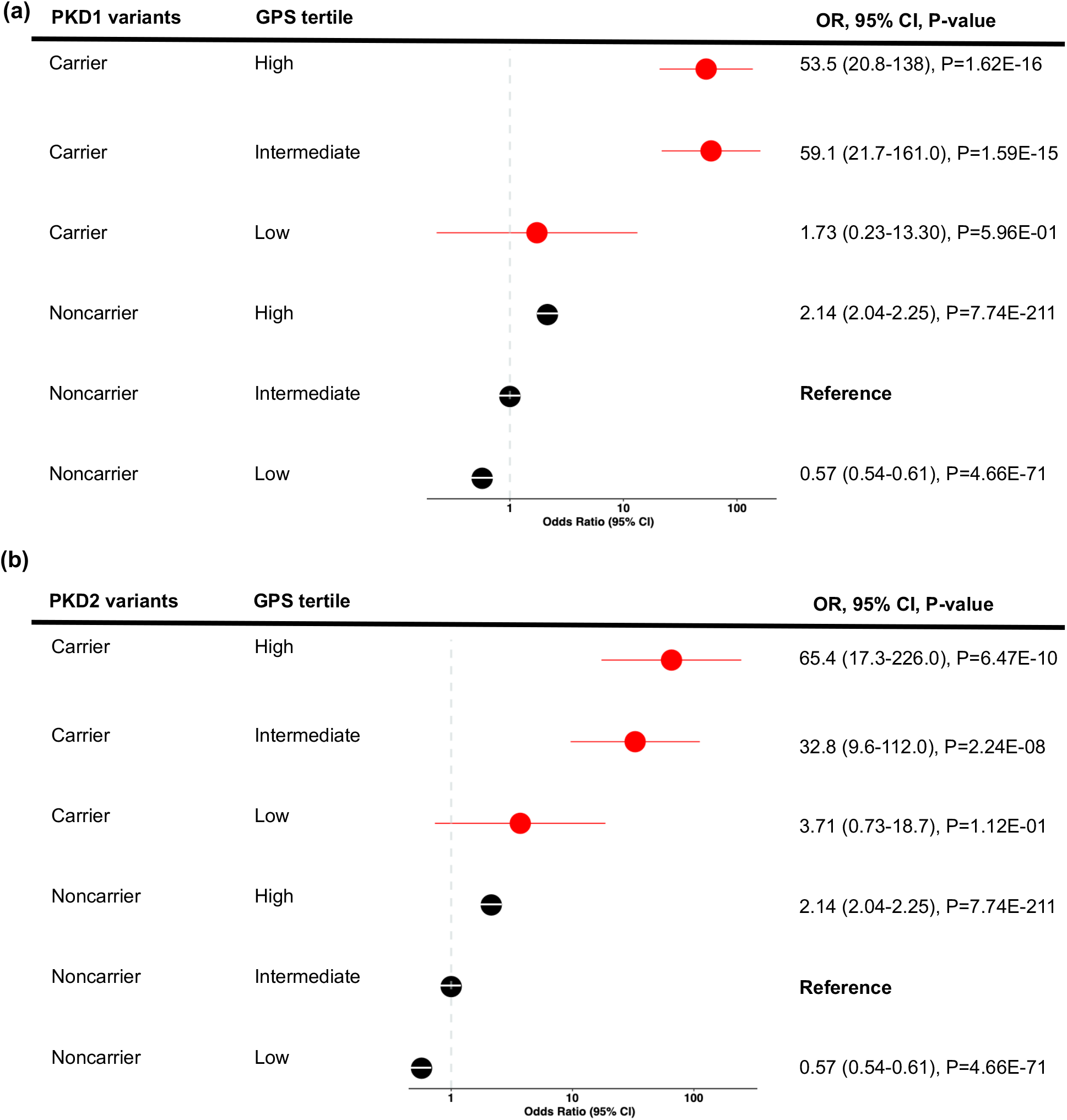
Polygenic effects on the risk of CKD in M1 variant carriers for **(a)** *PKD1* (N_total=_109) and **(b)** *PKD2* (N_total=_63) genes analyzed individually. Each polygenic risk score tertile for carriers was compared to the middle tertile of non-carriers (average population risk). X-axis shows Odds Ratios, the dotted vertical line corresponds to the OR=1.0 (no change in risk compared to the reference). Two-sided P-values derived from fixed effects meta-analysis are not adjusted for multiple testing. CI: Confidence Intervals.

**Supplementary Figure 4.**
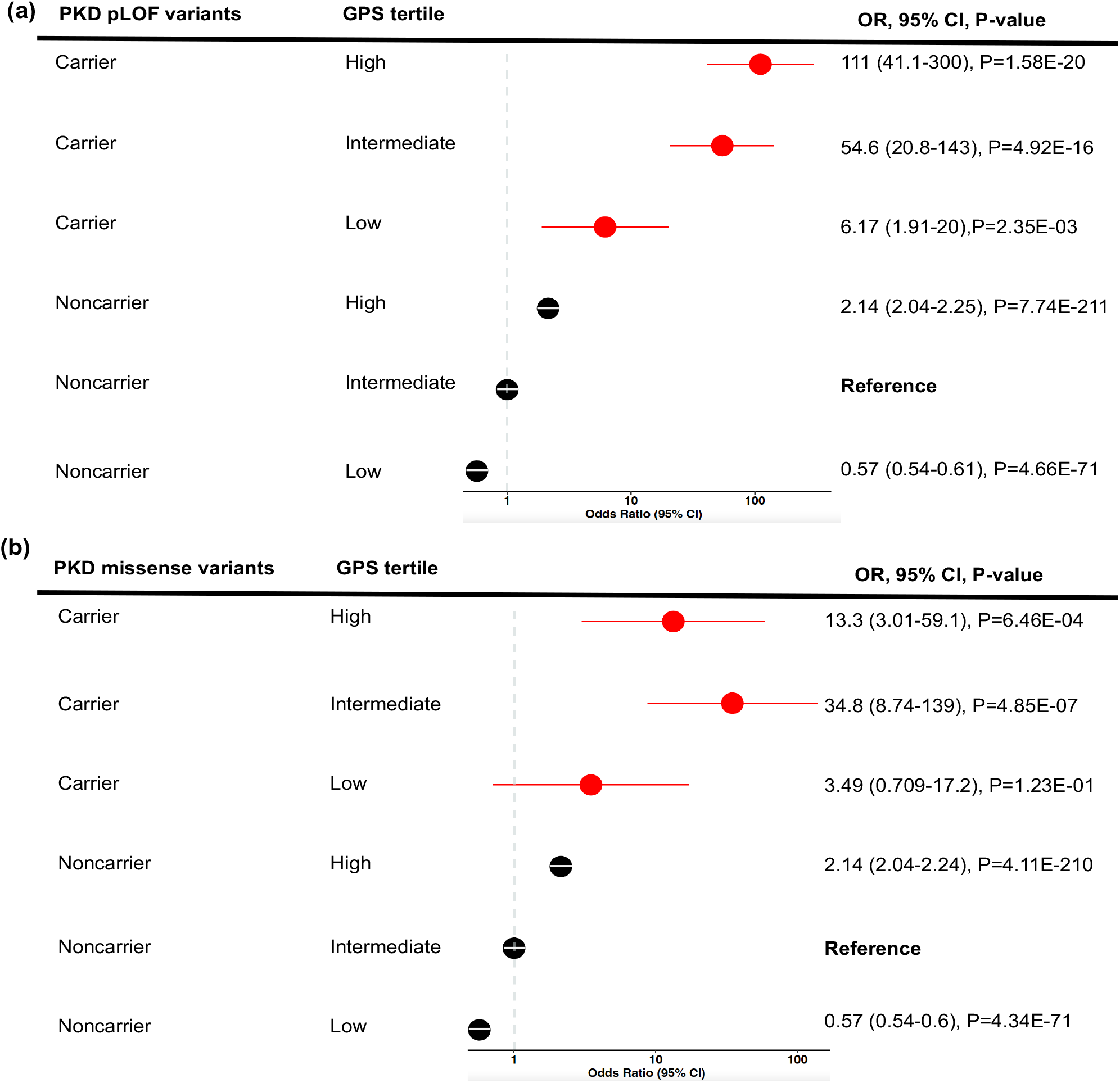
Polygenic effects on the risk of CKD in M1 variant carriers for **(a)** ADPKD pLOF variants (N_total=_111) and **(b)** ADPKD missense variants (N_total=_47) were analyzed individually. Each polygenic risk score tertile for carriers was compared to the middle tertile of non-carriers (average population risk). X-axis shows Odds Ratios, the dotted vertical line corresponds to the OR=1.0 (no change in risk compared to the reference). Two-sided P-values derived from fixed effects meta-analysis are not adjusted for multiple testing. CI: Confidence Intervals.

#### Supplementary Tables

**Supplementary Table 1.** Imputation of the AoU dataset using phase 3 1000 Genomes project reference panel (all populations): numbers of variants per chromosome before and after imputation by minor allelic frequency (MAF) and imputation quality (R2).

| Chromosomes | Array SNPs | Imputed SNPs<br>(MAF≥0.00 and<br>R2≥0) | Imputed SNPs<br>(MAF=0.001 and<br>R2=0.30) | Imputed SNPs<br>(MAF=0.001 and<br>R2=0.80) | Imputed SNPs<br>(MAF=0.01 and<br>R2=0.80) |
| --- | --- | --- | --- | --- | --- |
| 1 | 89688 | 3738240 | 2040921 | 1509280 | 909349 |
| 2 | 99772 | 4057613 | 2231592 | 1704713 | 1003370 |
| 3 | 84384 | 3355939 | 1879863 | 1455774 | 863987 |
| 4 | 76774 | 3338265 | 1899659 | 1474223 | 876432 |
| 5 | 69273 | 3032422 | 1704339 | 1320209 | 766074 |
| 6 | 82492 | 2954410 | 1694019 | 1328952 | 799683 |
| 7 | 63934 | 2753497 | 1539955 | 1158989 | 694545 |
| 8 | 60000 | 2651561 | 1470694 | 1135036 | 669886 |
| 9 | 49556 | 2063096 | 1121854 | 842974 | 505766 |
| 10 | 56241 | 2334090 | 1296803 | 986085 | 595717 |
| 11 | 56476 | 2333242 | 1286023 | 978430 | 581944 |
| 12 | 53025 | 2242720 | 1256253 | 945606 | 572855 |
| 13 | 40542 | 1661700 | 944885 | 726461 | 434997 |
| 14 | 36097 | 1535592 | 841576 | 636776 | 383314 |
| 15 | 35262 | 1404164 | 757414 | 555375 | 335228 |
| 16 | 38698 | 1549316 | 822821 | 591403 | 358495 |
| 17 | 33636 | 1345835 | 721632 | 495211 | 307640 |
| 18 | 33408 | 1319629 | 739218 | 551084 | 334086 |
| 19 | 24882 | 1084535 | 585112 | 379198 | 247001 |
| 20 | 27606 | 1047613 | 577463 | 420925 | 258119 |
| 21 | 16391 | 653791 | 353514 | 253922 | 159090 |
| 22 | 16388 | 652195 | 343797 | 234264 | 148765 |
| **Total** | **1,144,525** | **43,371,225** | **26,109,407** | **19,684,890** | **11,806,343** |

**Supplementary Table 2:** Genetic ancestry of the AoU dataset based on supervised machine learning with labeled phase 3 1000 Genomes project reference panel for training.

| Ancestry | N (%) | Age<br>(Mean in years) | Sex<br>(% Female) |
| --- | --- | --- | --- |
| EUR (European) | 94,376 (57) | 58.82 | 60.03 |
| AFR (African) | 36,380 (22) | 51.90 | 57.68 |
| AMR (Admixed American) | 28,807 (17) | 47.73 | 67.89 |
| EAS (East Asian) | 3,940 (2) | 47.04 | 63.76 |
| SAS (South Asian) | 1,705 (1) | 45.76 | 50.35 |
| **Total** | **165,208** | **50.25** | **59.94** |

**Supplementary Table 3:** Overall frequencies of *APOL1* G1 and G2 risk alleles and risk genotypes by ancestry and cohort.

| Dataset | Ancestry | APOL1- 1072A>G<br>(rs73885319) | APOL1- 1200T>G<br>(rs60910145) | APOL1-1212- del6<br>(rs71785313) | APOL1 risk<br>genotype (G1G1,<br>G1G2, or G2G2) |
| --- | --- | --- | --- | --- | --- |
| **UKBB** |  |  |  |  |  |
|  | African (AFR) | 0.28 | 0.28 | 0.15 | 0.12 |
|  | Europeans (EUR) | 3.75E-05 | 1.96E-04 | 2.91E-05 | <0.01 |
| **AoU** |  |  |  |  |  |
|  | African (AFR) | 0.233 | 0.232 | 0.139 | 0.12 |
|  | Admixed American (AMR) | 0.022 | 0.022 | 0.018 | 0.003 |
|  | Europeans (EUR) | 7.58E-04 | 7.58E-04 | 5.19E-04 | <0.01 |

**Supplementary Table 4:** The effect of ADPKD qualifying variant (QV) carrier status on the risk of CKD in the UK Biobank and the All of Us datasets. The ORs were adjusted for age, sex, diabetes, PCs of ancestry, and genotyping array or batch; the two-sided P-values correspond to the association tests of carrier status as a predictor of CKD and are not corrected for multiple testing; M1 includes only pLOF, and ‘P’ variants (N_total=_122 carriers); M2 includes pLOF, ‘P’, and ‘LP’ variants (N_total=_139 carriers); M3 includes pLOF and all deleterious missense variants as defined by 5 prediction algorithms, Revel >0.7, and not previously classified as ‘B’ or ‘LB’ by ClinVar (N_total=_267 carriers). All comparisons are made in reference to the common group of non-carriers (N_total=_297,539). CI: Confidence Intervals.

|  | ADPKD M1 carriers<br>OR (95%CI), P | ADPKD M2 carriers<br>OR (95%CI), P | ADPKD M3 carriers<br>OR (95%CI), P |
| --- | --- | --- | --- |
| **UKBB** | 18.2 (11.5-28.6), P=5.1E-36 | 17.3 (11.1-27.0), P=4.8E-36 | 7.1 (4.95-10.2), P=1.8E-26 |
| **AoU** | 8.36 (1.84-37.4), P=5.9E-03 | 6.11 (1.47-25.4), P=1.3E-02 | 3.13 (0.70-14.1), P=1.4E-01 |
| **Meta-analysis** | 17.1 (11.1-26.4), P=1.8E-37 | 15.8 (10.3-24.2), P=5.2E-37 | 6.77 (4.76-9.60), P=1.0E-26 |

**Supplementary Table 5:**
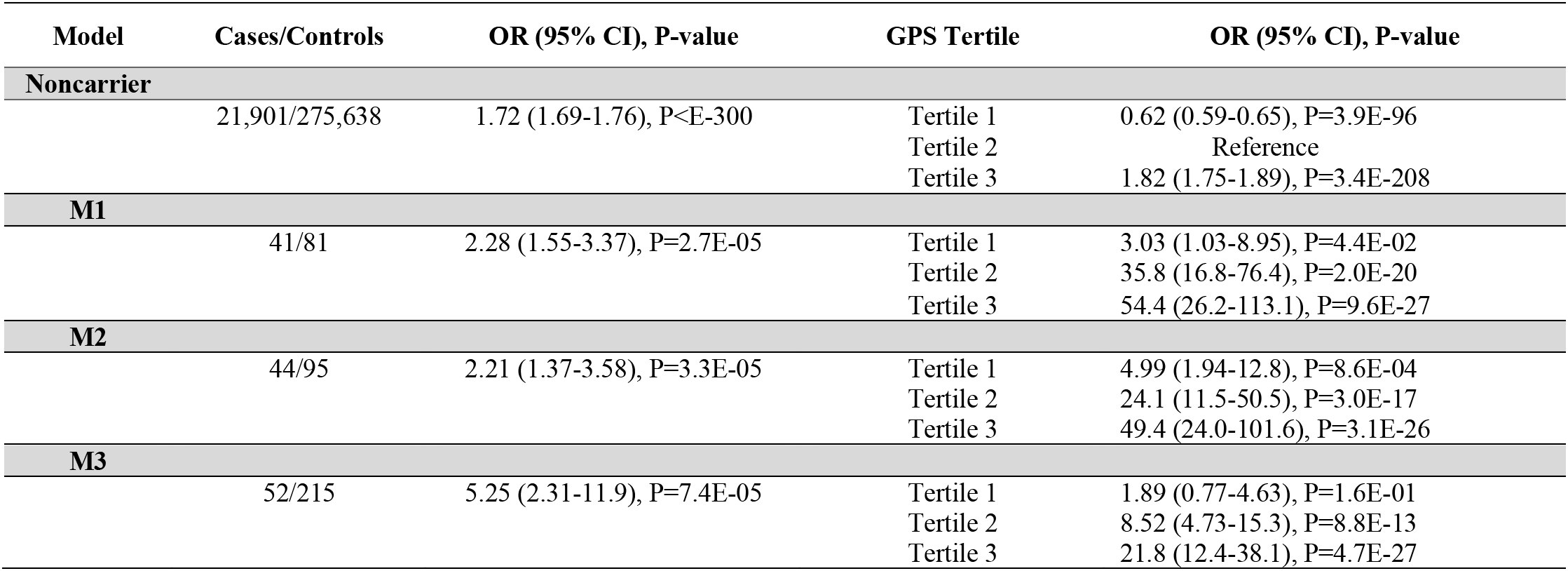
Effects of GPS on the risk of CKD in the ADPKD M1, M2, and M3 variant carriers in the meta-analysis of UKBB and AoU. All effect estimates were calculated in reference to the middle tertile of non-carriers (average risk) and were adjusted for age, sex, diabetes, PCs of ancestry, and genotyping array or batches. Two-sided P-values correspond to fixed effects meta-analysis and are not corrected for multiple testing.

**Supplementary Table 6:** The effect of COL4A-AN qualifying variant (QV) carrier status on the risk of CKD in the UK Biobank and the All of Us datasets. The ORs were adjusted for age, sex, diabetes, PCs of ancestry, and genotyping array or batch; the two-sided P-values correspond to the association tests of carrier status as a predictor of CKD and are not corrected for multiple testing; M1 includes only pLOF, and ‘P’ variants (N_total=_1,292 carriers); M2 includes pLOF, ‘P’, and ‘LP’ variants (N_total=_1,458 carriers); M3 includes pLOF and all deleterious missense variants as defined by 5 prediction algorithms, Revel >0.7, and not previously classified as ‘B’ or ‘LB’ by ClinVar (N_total=_2,056 carriers); M3 recessive model (N_total=_127) includes biallelic carriers of M3 variants for COL4A3 or COL4A4, or M3 hemizygous males. All comparisons are made in reference to the common group of non-carriers (N_total=_298,778).

| Datasets | COL4A-AN M1 carriers<br>OR (95%CI), P | COL4A-AN M2 carriers<br>OR (95%CI), P | COL4A-AN M3 carriers<br>OR (95%CI), P | COL4A-AN M3 recessive<br>OR (95%CI), P |
| --- | --- | --- | --- | --- |
| UKBB | 1.41 (1.15-1.74), P=1.1E-03 | 1.34 (1.03-1.73), P=3.0E-02 | 1.55 (1.26-1.92), P=5.0E-05 | 3.10 (1.66-5.78), P=4.2E-04 |
| AoU | 1.21 (0.82-1.79), P=3.3E-01 | 1.05 (0.67-1.61), P=8.2E-01 | 1.29 (0.90-1.85), P=1.6E-01 | 6.69 (1.23-36.4), P=2.8E-02 |
| Meta | 1.37 (1.13-1.64), P=8.5E-04 | 1.25 (1.00-1.56), P=4.9E-02 | 1.48 (1.23- 1.77), P=2.6E-05 | 3.38 (1.88-6.08), P=4.7E-05 |

**Supplementary Table 7:** Effects of GPS in the COL4-AN M1, M2, and M3 variant carriers in the meta-analysis of UKBB and AoU. All effect estimates were calculated in reference to the middle tertile of non-carriers (average risk) and were adjusted for age, sex, diabetes, PCs of ancestry, and genotyping array or batches. Two-sided P-values correspond to fixed effects meta-analysis and are not corrected for multiple testing.

| Model | Cases/Controls | OR (95% CI), P-value | GPS Tertile | OR (95% CI), P-value |
| --- | --- | --- | --- | --- |
| **Noncarrier** |  |  |  |  |
|  | 21,446/277,332 | 1.70 (1.68-1.73), P<E-300 | Tertile 1 | 0.61 (0.59-0.64), P=2.5E-98 |
|  |  |  | Tertile 2 | Reference |
|  |  |  | Tertile 3 | 1.82 (1.75-1.89), P=8.1E-210 |
| **M1** |  |  |  |  |
|  | 99/1,193 | 1.78 (1.22-2.58), P=2.4E-03 | Tertile 1 | 1.08 (0.63-1.86), P=7.7E-01 |
|  |  |  | Tertile 2 | 1.66 (1.03-2.68), P=3.7E-02 |
|  |  |  | Tertile 3 | 2.53 (1.66-3.85), P=1.4E-05 |
| **M2** |  |  |  |  |
|  | 112/1,344 | 2.47 (1.56-3.94), P=1.3E-04 | Tertile 1 | 0.66 (0.40-1.07), P=9.6E-02 |
|  |  |  | Tertile 2 | 1.26 (0.84-1.88), P=5.3E-01 |
|  |  |  | Tertile 3 | 2.55 (1.83-3.56), P=3.1E-08 |
| **M3** |  |  |  |  |
|  | 172/1,884 | 1.93 (1.26-2.95), P=2.3E-03 | Tertile 1 | 1.10 (0.57-2.12), P=7.7E-01 |
|  |  |  | Tertile 2 | 1.45 (0.82-2.55), P=2.0E-01 |
|  |  |  | Tertile 3 | 2.77 (1.73-4.46), P=2.7E-05 |
| **M3 (recessive)** |  |  |  |  |
|  | 21/106 | 1.19 (0.69-2.07), P=5.2E-01 | Tertile 1 | 2.29 (0.64-8.12), P=2.0E-01 |
|  |  |  | Tertile 2 | 2.71 (0.97-7.59), P=5.6E-02 |
|  |  |  | Tertile 3 | 6.73 (2.59-17.5), P=8.8E-05 |

**Supplementary Table 8:** GPS effect estimates for each *COL4A* gene under the M1 model. Only UKBB data included due to low case counts in the AoU dataset. The P-values are two-sided and not adjusted for multiple testing.

| Gene | Cases/controls | OR (95% CI), P-value | GPS Tertile | OR (95% CI), P-value |
| --- | --- | --- | --- | --- |
| ***COL4A3*** |  |  |  |  |
|  | 12/211 | 1.37 (0.60-3.14), P=4.5E-01 | Tertile 1 | 1.70 (0.65-4.43), P=2.8E-01 |
|  |  |  | Tertile 2 | 1.20 (0.36-3.98), P=7.7E-01 |
|  |  |  | Tertile 3 | 2.00 (0.70-5.72), P=1.9E-01 |
| ***COL4A4*** |  |  |  |  |
|  | 16/313 | 1.49 (0.73-3.03), P=2.7E-01 | Tertile 1 | 0.86 (0.264-2.78), P=7.9E-01 |
|  |  |  | Tertile 2 | 1.69 (0.70-4.05), P=2.3E-01 |
|  |  |  | Tertile 3 | 2.23 (0.98-5.06), P=5.6E-02 |
| ***COL4A3 or 4*** |  |  |  |  |
|  | 28/524 | 1.91 (0.90-4.09), P=9.3E-02 | Tertile 1 | 0.94 (0.41-2.18), P=8.8E-01 |
|  |  |  | Tertile 2 | 1.54 (0.78-3.02), P=2.1E-01 |
|  |  |  | Tertile 3 | 2.43 (1.31-4.52), P=5.0E-03 |
| ***COL4A5*** |  |  |  |  |
|  | 3/32 | 1.97 (0.18-21.10), P=5.7E-01 | Tertile 1 | 3.46 (0.40-29.8), P=2.5E-01 |
|  |  |  | Tertile 2 | 1.35E-04 (2.6E-82-7.0E73), P=9.2E-01 |
|  |  |  | Tertile 3 | 11.0 (2.02-60.2), P=5.6E-03 |

